# Diagnosis of Metabolic Syndrome using Machine Learning, Statistical and Risk Quantification Techniques: A Systematic Literature Review

**DOI:** 10.1101/2020.06.01.20119339

**Authors:** Habeebah Adamu Kakudi, Chu Kiong Loo, Foong Ming Moy

## Abstract

Metabolic syndrome (MetS), known to substantially lower the quality of life is associated with the increased incidence of non-communicable diseases (NCDs) such as type II diabetes mellitus, cardiovascular diseases and cancer. Evidence suggests that MetS accounts for the highest global mortality rate. For the early and accurate diagnosis of MetS, various statistical and ML techniques have been developed to support its clinical diagnosis. We performed a systematic review to investigate the various statistical and machine learning techniques (ML) that have been used to support the clinical diagnoses of MetS from the earliest studies to January 2020. Published literature relating to statistical and ML techniques for the diagnosis of MetS were identified by searching five major scientific databases: PubMed, Science Direct, IEEE Xplore, ACM digital library, and SpringerLink. Fifty-three primary studies that met the inclusion criteria were obtained after screening titles, abstracts and full text. Three main types of techniques were identified: statistical (n = 10), ML (n = 40), and risk quantification (n = 3). Standardized Z-score is the only statistical technique identified while the ML techniques include principal component analysis, confirmatory factory analysis, artificial neural networks, multiple logistics regression, decision trees, support vector machines, random forests, and Bayesian networks. The areal similarity degree risk quantification, framingham risk score and simScore were the three risk quantification techniques identified. Evidence suggests that evaluated ML techniques, with accuracy ranging from 75.5% to 98.9%, can more accurately diagnose MetS than both statistical and risk quantification techniques. The standardised Z-score is the most frequent statistical technique identified. However, highlighted proof based on performance measures indicate that the decision tree and artificial neural network ML techniques have the highest predictive performance for the prediction of MetS. Evidence suggests that more accurate diagnosis of MetS is required to evaluate the predictive performance of the statistical and ML techniques.

---

Version June 1, 2020 submitted to Appl. Sci.

## 1 Introduction

Rapid urbanisation, excess energy intake, increased obesity and prevalent sedentary lifestyle habits are majory responsible for the steady increase of metabolic syndrome (Mets) and its associated diseases [1]. MetS is a cluster of metabolic abnormalities mainly hyperinsulinemia (insulin resistance), central obesity (raised waist circumference), dyslipidemia (increase in triglyceride and decrease in high-density lipoprotein cholesterol), hypertension (increase in blood pressure), and microalbuminuria [2–4]. The definition of MetS has been iteratively changed and improved by numerous health expert groups to cater for various population needs and disease trends globally. These health expert groups include The European Group for the Study of Insulin Resistance (EGIR) [4], the National Cholesterol Education Program —Third Adult Treatment Panel (NCEP ATP III) [3], the American Association of Clinical Endocrinologists, American Heart Association— National Heart, Lung, and Blood Institute (AHA/NHLBI) [5], the International Diabetes Federation (IDF) [6], and the Joint Interim Statement (JIS) [7]. Table 1 presents the commonly accepted and clinically adopted definitions of MetS based on the five (5) MetS risk factors (MetSR-F) common to all the clinical definitions: Fasting Plasma Glucose (FPG), Waist Circumference (WC), Triglycerides (TG), High-Density Lipoprotein Cholesterol (HDL-C), and Blood Pressure (BP). The most up-to-date version is the dichotomous definition in 2009 by the JIS [7] which states existence of MetS provided that at least three out of the following five risk factor abnormalities are present:

1. Central obesity (ethnic- and country- specific);
2. Raised triglyceride: TG ≥ 150 mg/dL(1.7 mmol/L);
3. Reduced high-density lipoprotein: HDL-C < 50 mg/dL (1.3 mmol/L) in women and HDL-C <40 mg/dL (1.0 mmol/L) in men;
4. Increased blood pressure (BP): - systolic blood pressure *≥* 130 mm Hg and/or diastolic blood pressure *≥* 85 mm Hg; and
5. Elevated fasting plasma glucose: FPG *≥* 100 mg/dL (5.6 mmol/L).

**Table 1.**
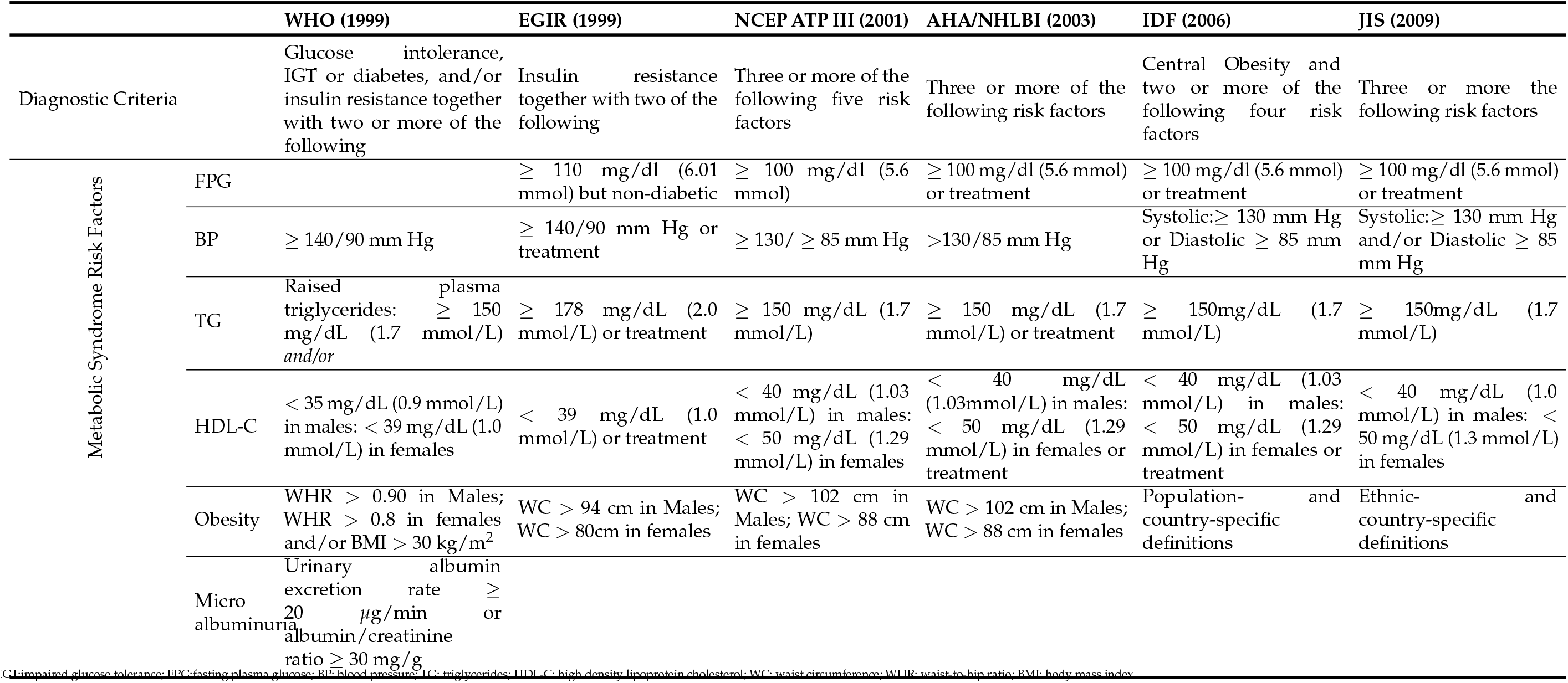
Clinical definitions of metabolic syndrome

## 2 Challenges and Motivation

MetS is a growing epidemic and a major global socio-economic burden [8]. MetS has been ascertained to be an active biomarker of three major non-communicable diseases (NCDs): cardiovascular diseases (CVDs), type II diabetes mellitus (T2DM), and cancer [8–10]. Other diseases known to be associated with MetS include osteoporosis [11], kidney stone [12], upper gastrointestinal disease [13], Psoriasis [14], alcohol use disorders [15], and anxiety [16].

CVDs, T2DM, and cancer are responsible for the motility of 17.7 million, 1.6 million, and 8.8 million people respectively yearly. Thus, constituting major health-care burdens around the world with the highest level of mortality rate. Furthermore, the global prevalence of MetS is such that it affects one out of every three persons in the United States [17,18], a quarter of the European population[19], and one-sixth of the Asian population [20].

In a bid to further understand current researches relating to MetS, existing literature reviews have been conducted with a focus on answering several research questions relating to the characteristics and associated diseases of MetS.

Timar and colleagues, [1] and Lopez-Candales [21] presented a summary of MetS by reviewing each MetSR-F, cut-off thresholds, and individuals that are most susceptible to the abnormalities. The review by Palomo and colleagues, [22] showed that MetS is characterised by alterations in hemostatis and fibrinolysis which was attributed to the metabolic abnormalities. In their survey, Gami and coworkers, [23] found thirty-seven studies that ascertained the association between MetS and CVDs. They concluded that individuals who present with the risk of MetS are highly at risk of developing CVDs which could increase mortality rate if lifestyle and preventive interventions are not applied. Xue and Michels, [9] reviewed available evidence that proved the existence of a clear relation between MetS, T2DM, and the onset of breast cancer.

Recently, Wong and colleagues, [12] noted a definite association among breast cancer, kidney stone and MetS caused by the presence of abnormalities such as obesity, hypertension, hyperinsulinemia and insulin resistance. Another study [24] noticed the increased risk of developing MetS in patients treated with antipsychotic agents due to sedentary lifestyle, unhealthy food choices and high rate of smoking. These factors subsequently led to weight gain, and increased prevalence of diabetes and CVDs. Motillo et al. [25] found that MetS is related to a 2-times increase in CVD and a 1.5 times increase in overall mortality rate while Kaur et al. [26] in their extensive review summarised existing literature related to the definition of MetS, its epidemiology and intervention approaches.

In all, these aforementioned studies have only analysed the current state-of-the-art of MetS and some of its associated diseases; however, none of them have examined the existing statistical, machine learning and risk quantification techniques that support the clinical diagnosis of MetS. We consider that the analyses of research activity in this domain is of utmost importance in order to investigate further research possibilities aimed at early diagnosis and prevention of MetS.

The present systematic review is aimed to identify and assess machine learning (ML), statistical and risk quantification (RQ) techniques which support the clinical diagnosis of MetS. The justification behind this systematic review is based on the requirement of knowledge acquisition that could assist in improving the quality of non-clinical techniques for the early diagnosis of MetS and subsequently to promote the management of MetS in clinical practice.

## 3 Review Method

The systematic review method adopted in this paper follows the procedures given by the PRISMA guidelines [27]. Therefore, we developed the study protocol by designing the search strategy, enumerating inclusion/exclusion criteria, extracting, and synthesising extracted data. In the first step, we carried out the search strategy by identifying the search terms and the electronic database to carry out our search. Relevant primary studies were selected as depicted in the PRISMA flow chart in Fig 1.

**Figure 1.**
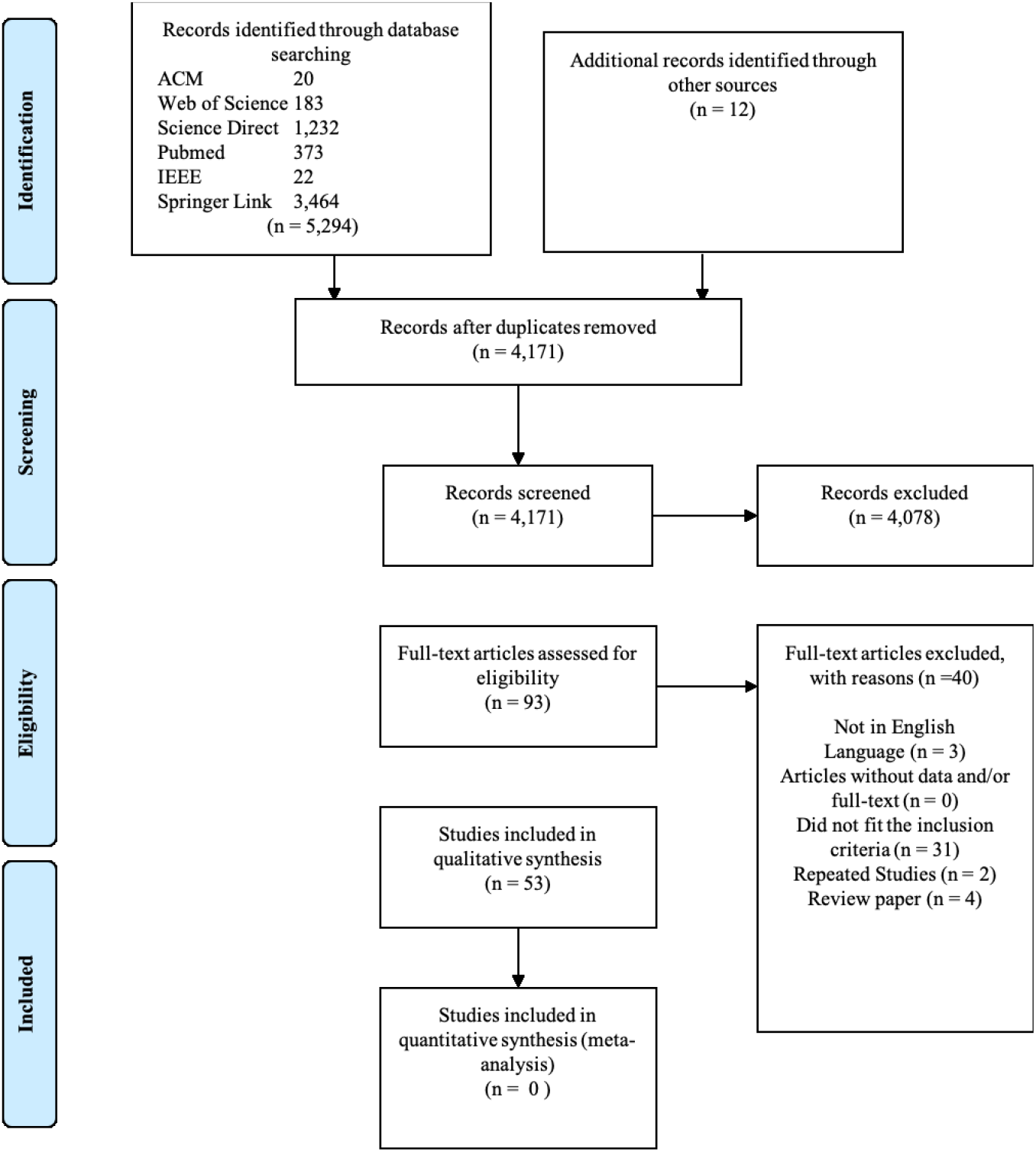
PRISMA flow chart for included and excluded studies in the systematic review on metabolic syndrome diagnosis methods

### 3.1 Search strategy

The first search was carried out from the earliest years until February 2020 in the following electronic data sources: Association of Computing Machinery (ACM) Digital Library, PubMed, Science Direct, Web of Science, and IEEE Xplore Digital Library. These electronic databases are widely accepted by various research communities [28].

A total of 54 search terms were used to collect the articles in this systematic review. These search terms cut across the different types of statistical and machine learning techniques that are most frequently used for classification and prediction. The main search term of the systematic review was “Metabolic Syndrome”. The second search term consisted of relevant machine learning keywords with “Metabolic Syndrome” using the ’AND’ and ’OR’ boolean concatenators as presented in Table∼2. However, the above mentioned search procedure in IEEE digital library revealed ambiguous search results of 504,794 publications even after refining to remove conference papers. A search into these results did not reveal any publication based on MetS. Furthermore, the same search conducted in ACM digital library revealed no results at all. Therefore only the main search term ’Metabolic Syndrome” was used in these two databases and this search revealed relevant results in both digital libraries. The whole search was carried out in the University of Malaya library electronic database.

**Table 2.** Search phrases and the number of articles found

| Search No. | Search strings | Sources and number of publications obtained |  |  |  |  |  |
| --- | --- | --- | --- | --- | --- | --- | --- |
|  |  | SD | WoS | ACM | PubMed | IEEE | Springer |
| 1 | ("Metabolic Syndrome") AND ("Naive Bayes" OR "Bayesian Network" OR "Decision tree" OR "Classification and regression tree" OR "Chi-squared Automatic Interaction Detection" OR "Linear discriminant analysis" OR "K-nearest neighbours algorithm") | 11 | 57 | 1 | 36 | 6 | 371 |
| 2 | ("Metabolic Syndrome") AND ("Learning vector quantization" OR "Self-organizing map" OR "K-means" OR "Expectation-maximization" OR "Hierarchical Clustering" OR "Fuzzy clustering" OR "Hierarchical clustering" OR "Hidden Markov models") | 11 | 36 | 2 | 26 | 2 | 546 |
| 3 | ("Metabolic Syndrome") AND ("Logistic regression" OR "Ordinary least squares regression" OR "Linear regression" OR "Stepwise regression" OR "AdaBoost" OR "Boosting" OR "Random Forest" OR "Gaussian mixture models" OR "k-Means clustering") | 1091 | 5 | 1 | 12 | 3 | 1196 |
| 4 | ("Metabolic Syndrome") AND ("machine learning" or "deep learning" OR "data mining" OR "predictive models" OR "Artificial neural network" OR "Back-Propagation" OR "Multilayer Perceptron" OR "association rule mining") | 7 | 14 | 10 | 73 | 9 | 860 |
| 5 | ("Metabolic Syndrome") AND ("Confirmatory factor analysis" OR "Factor analysis" OR "Principal component regression" OR "Principal Component Analysis") | 112 | 71 | 6 | 226 | 2 | 491 |
| Total |  | 1232 | 183 | 20 | 373 | 22 | 3464 |

### 3.2 Study selection

The whole initial search from the electronic databases revealed 6582 studies. These studies were screened based on the objective of the review which sought to appraise the empirical quality of the studies as well as evaluate the effectiveness of applying non clinical methods to diagnose MetS. After removing duplicates, the first selection step included screening the titles, abstracts, and keywords of the studies for eligibility in relation to the following inclusion criterion:

i. The study participants were humans with or without metabolic syndrome and the participant characteristics were clearly defined.
ii. The study objective involved the diagnosis of metabolic syndrome using a non clinical approach in addition to or as opposed to the clinical dichotomous definition.
iii. At least an outcome for the non-clinical diagnosis of metabolic syndrome exists in the study.

Ninety-three primary studies were left after the first screening. We were able to retrieve the full text of all 93 articles by searching online databases. The inclusion/exclusion criteria were independently tested by two reviewers (HAK and MFM) and a conclusion was agreed upon after comprehensive discussions. The entire full text of the 93 primary studies were screened by two reviewers (HAK and MFM) based on the following exclusion criteria:

i. Non-peer reviewed studies such as tutorials, reports, conference, and editorial papers.
ii. Studies with adolescent and children participants.
iii. Studies based on conceptual frameworks or structures without empirical analysis and results.

The two researchers compared their results and where discrepancies existed between two results of the same paper, a joint assessment was carried out with all the researchers and an agreement was made to either include or exclude the study from the systematic review. Finally, 40 studies were excluded and a total of 53 primary studies were identified for inclusion into this systematic review.

### 3.3 Data extraction

Reviewer HAK independently extracted data from the primary studies by filling a data extraction form and gathering general information from each study such as author name, publication year, title, study of dataset, country, study population, study design, MetS diagnosis techniques and evaluation metrics results. All data extracted is presented in Table 3 Researchers HAK and CKL verified the soundness of data by making sure that information extracted from each study justified the aim of the research.

**Table 3.**
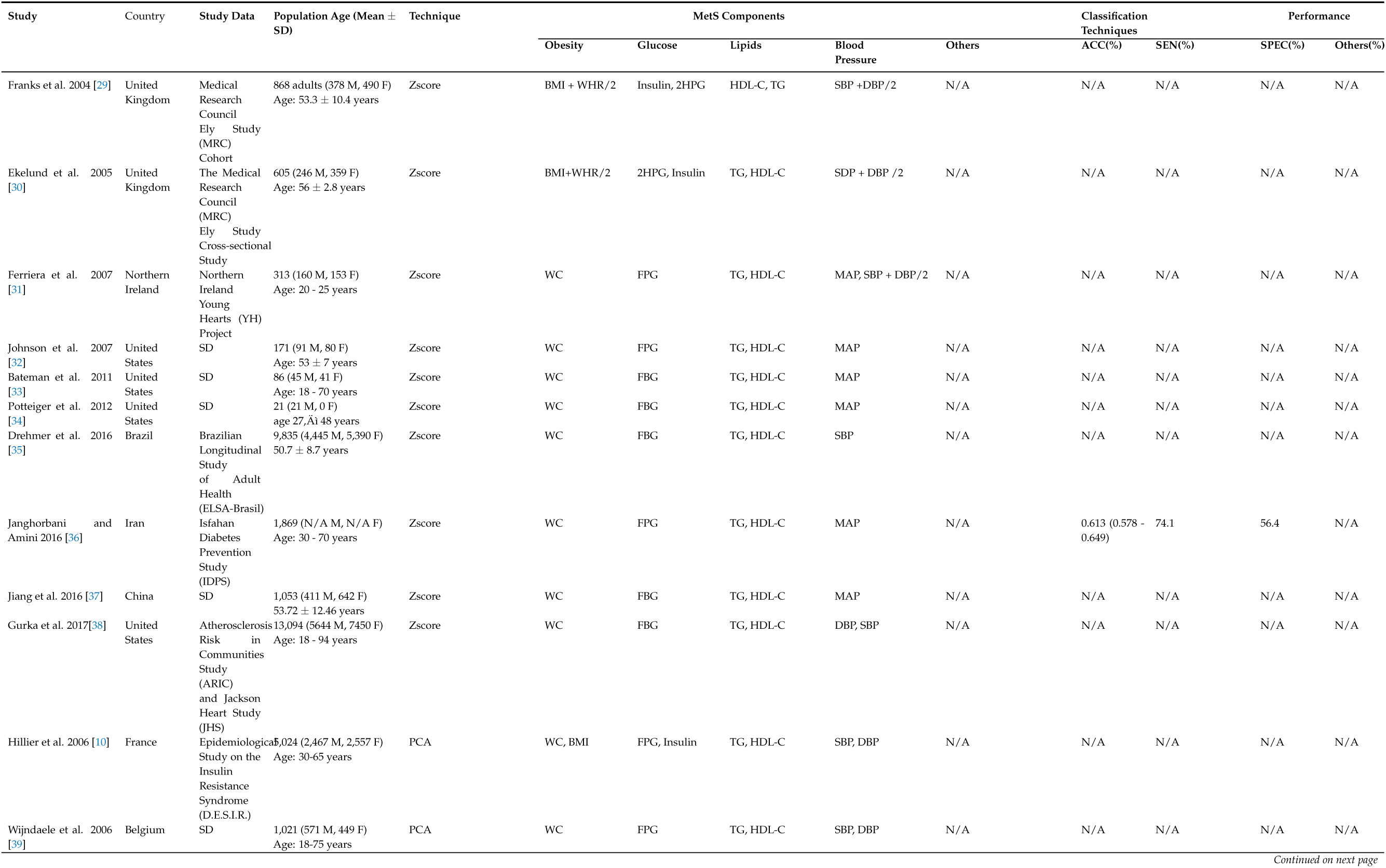
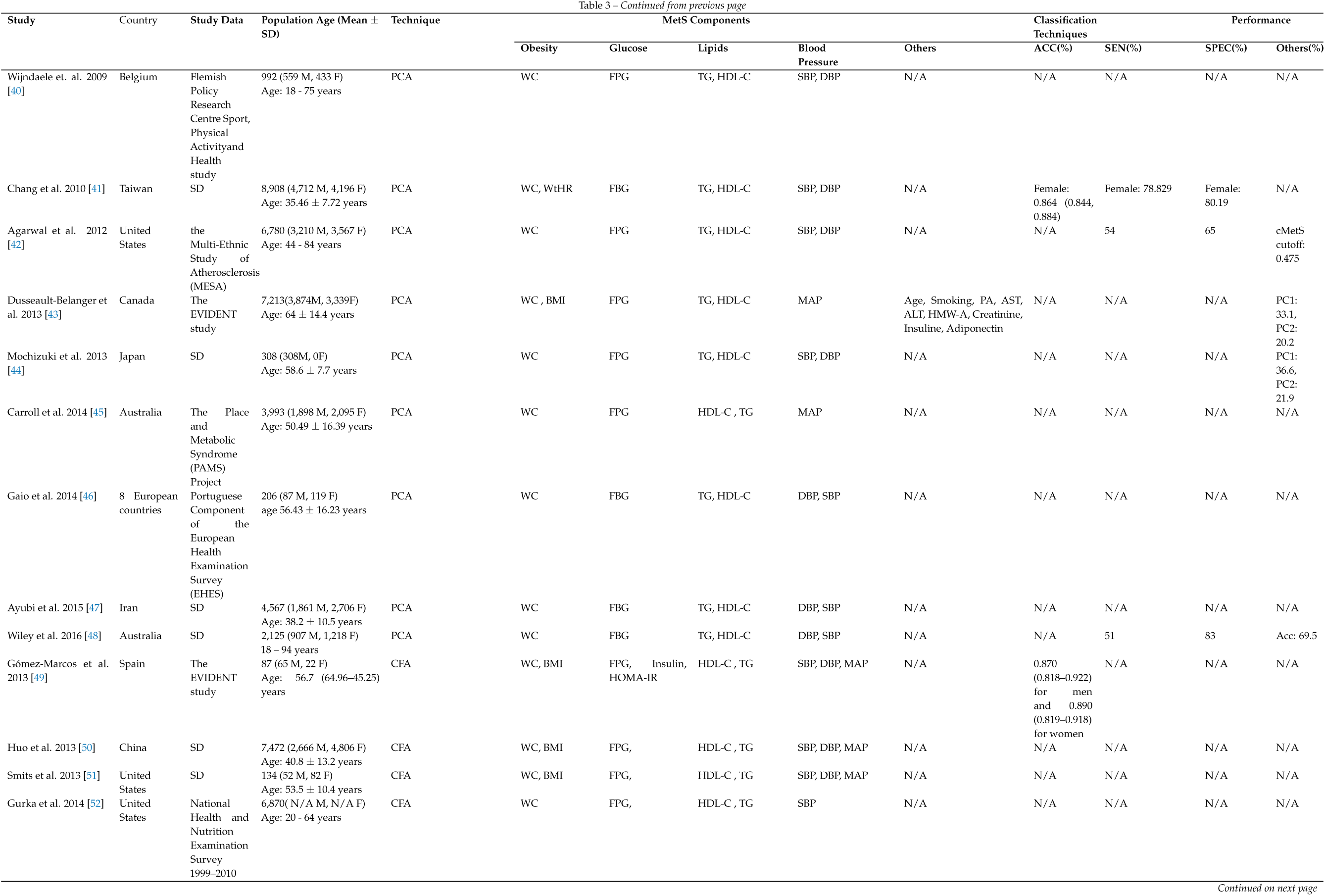
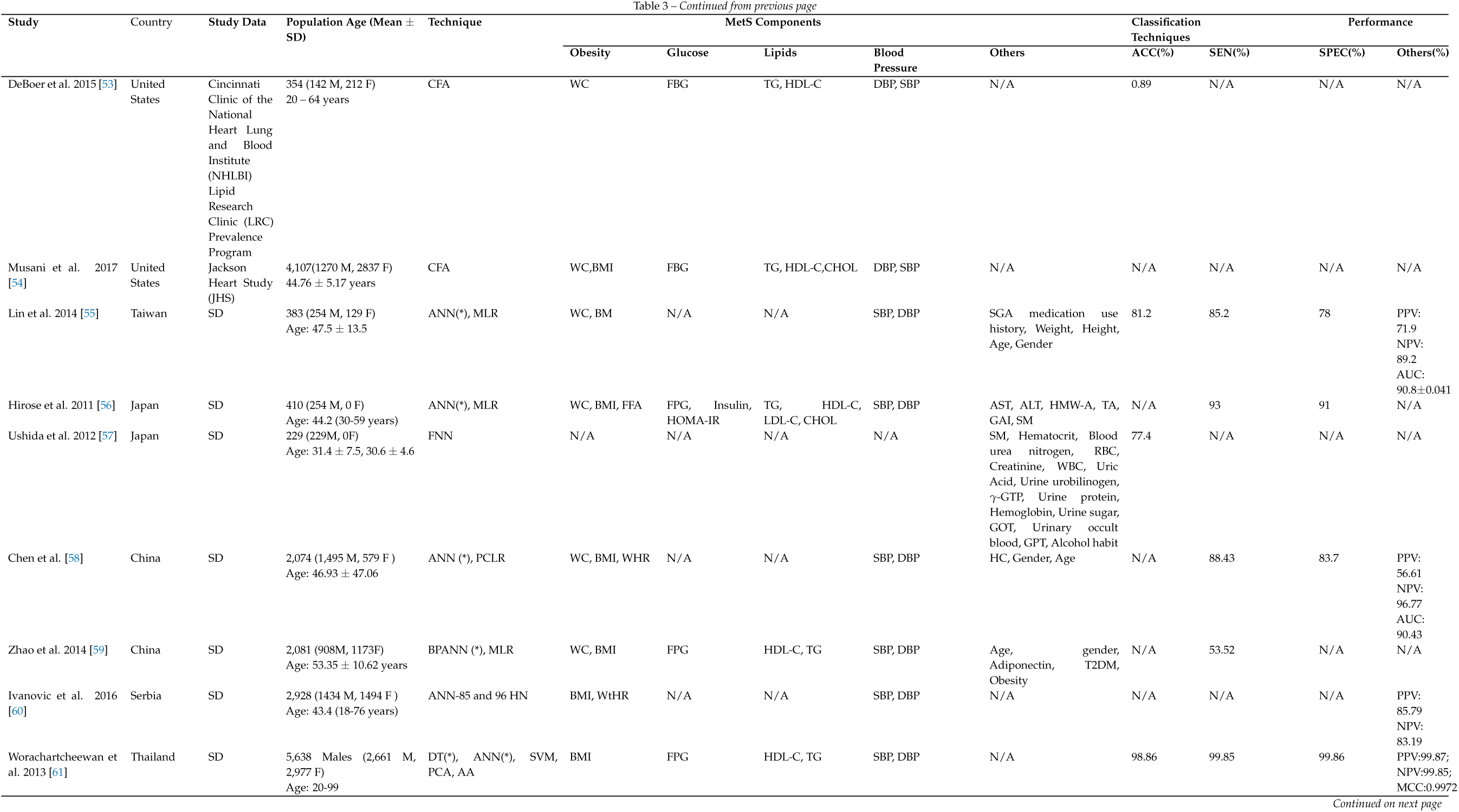
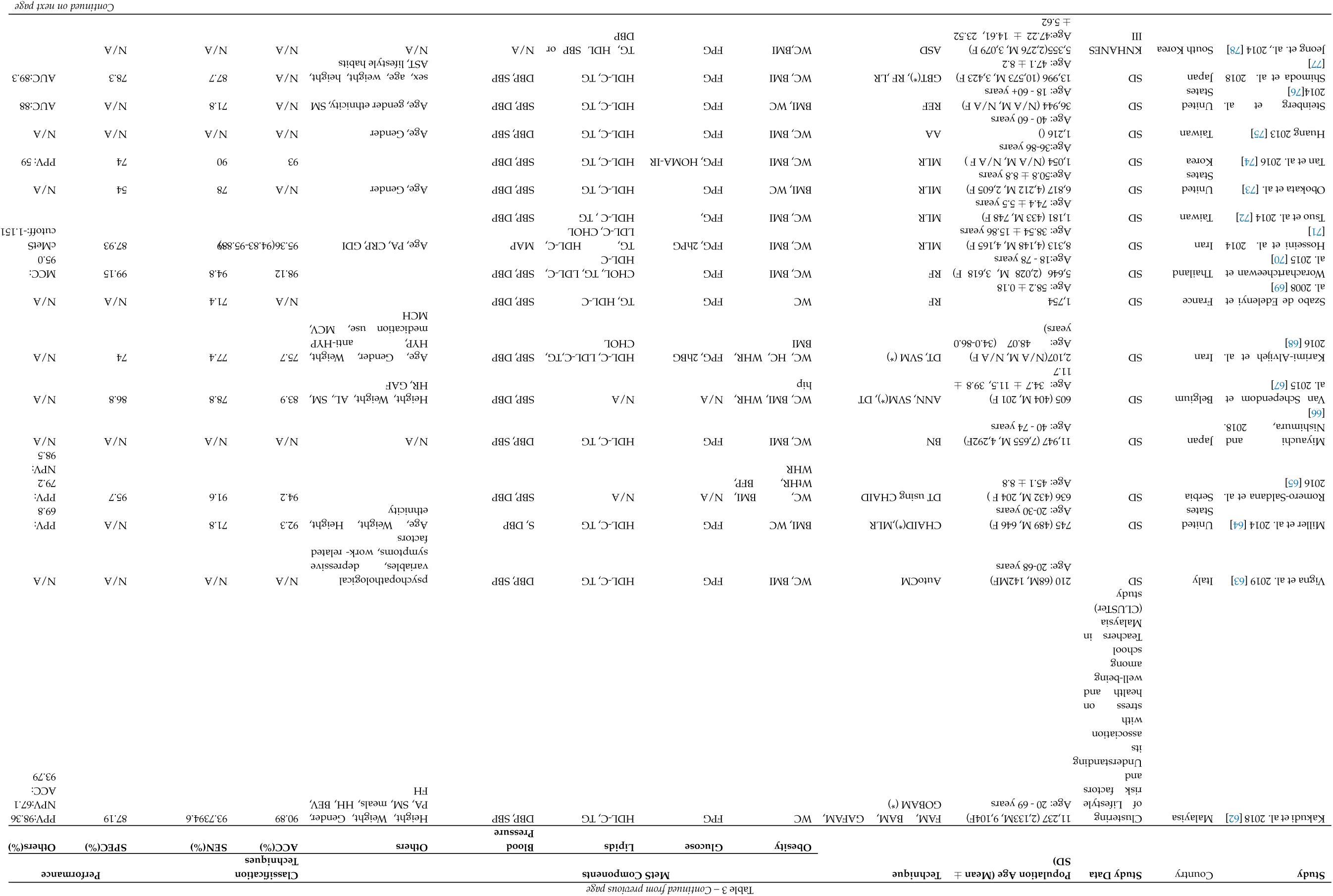
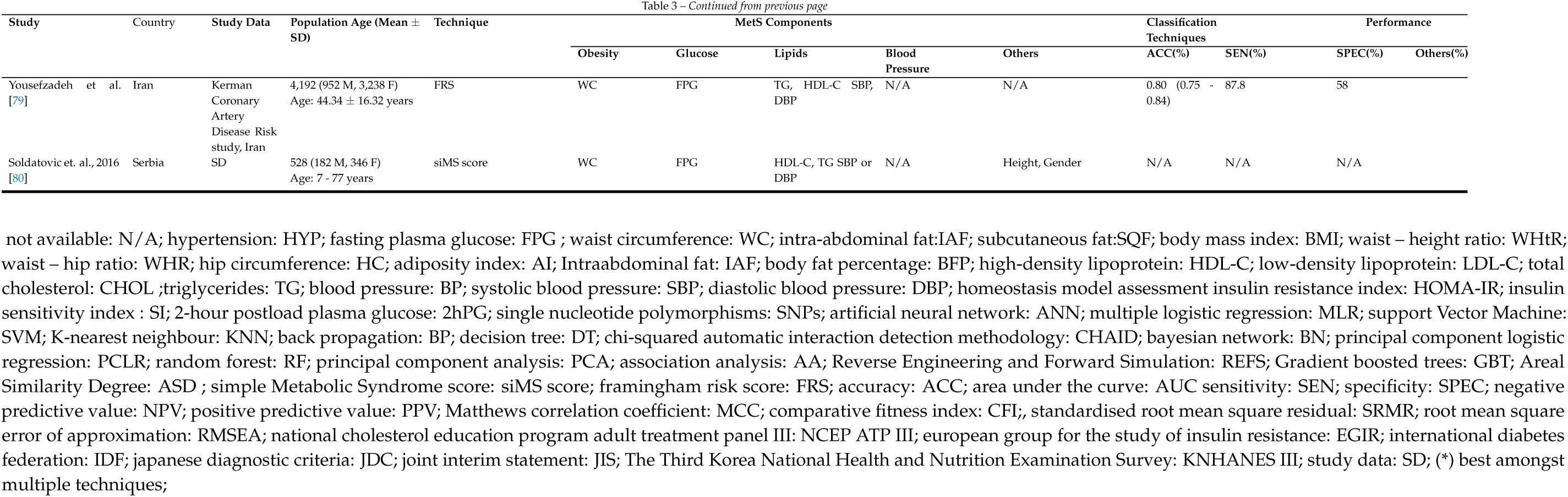
Metabolic syndrome risk orediction using ML, statistical and RQ techniques

#### 3.3.1 Publication Years

The search for this review was carried out from the earliest studies to January 2020. This is to identify all the non-clinical techniques for the diagnosis of MetS. From the distribution in Figure 2, the number of research on non-clinical methods peaked in the year 2014. The research interest trend has decreased from 2016 till date by 10%. Only one study was published in 2019, affirming the inclusion of up-to-date relevant studies in this systematic review.

**Figure 2.**
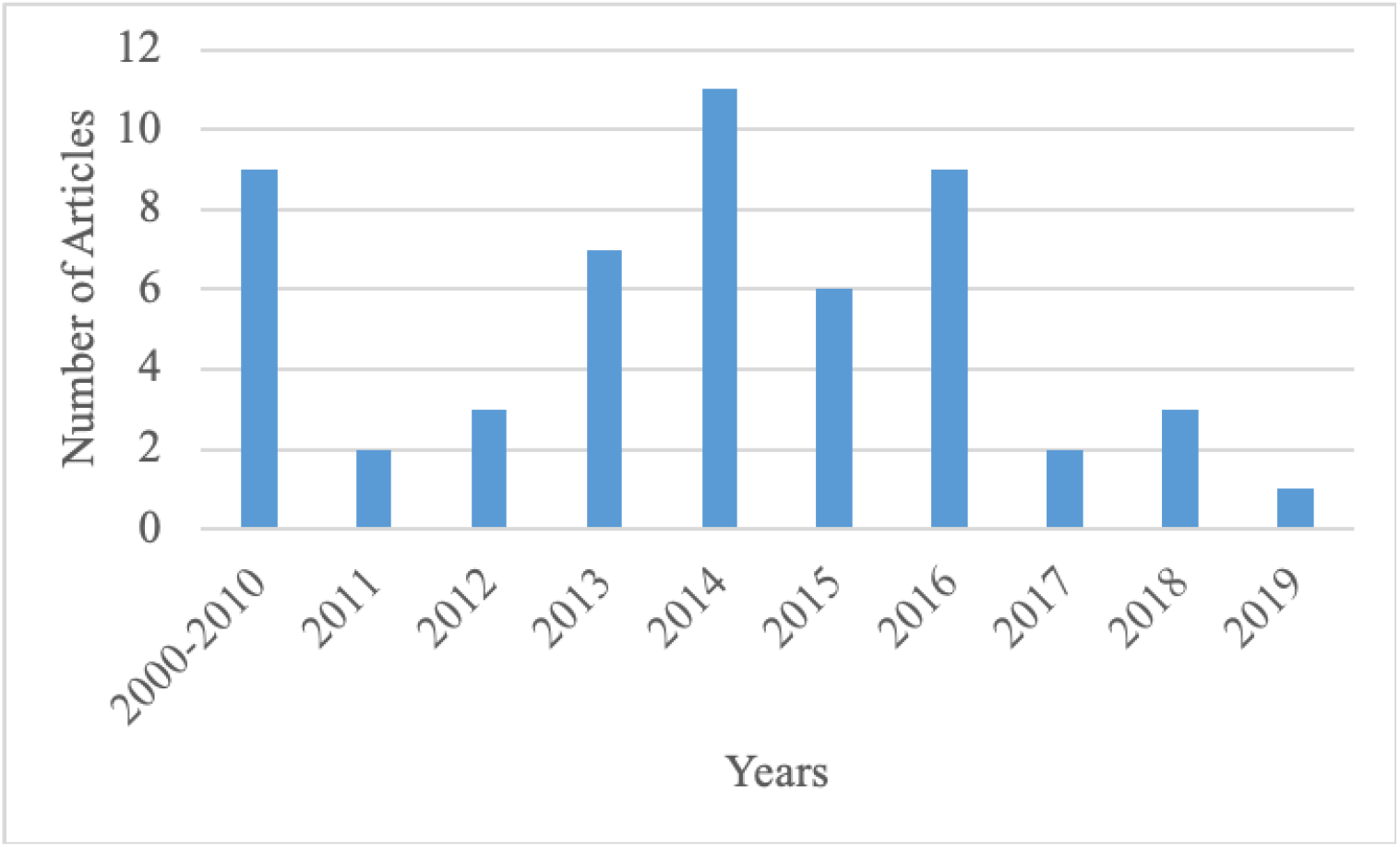
Publication years of primary studies

#### 3.3.2 Place of Study

The distribution of studies by the countries where their study data was collected is presented in Figure 3. The highest number of studies, 23%, were published from the United States. This was followed by Iran and Japan where each country published 10% of the studies.

**Figure 3.**
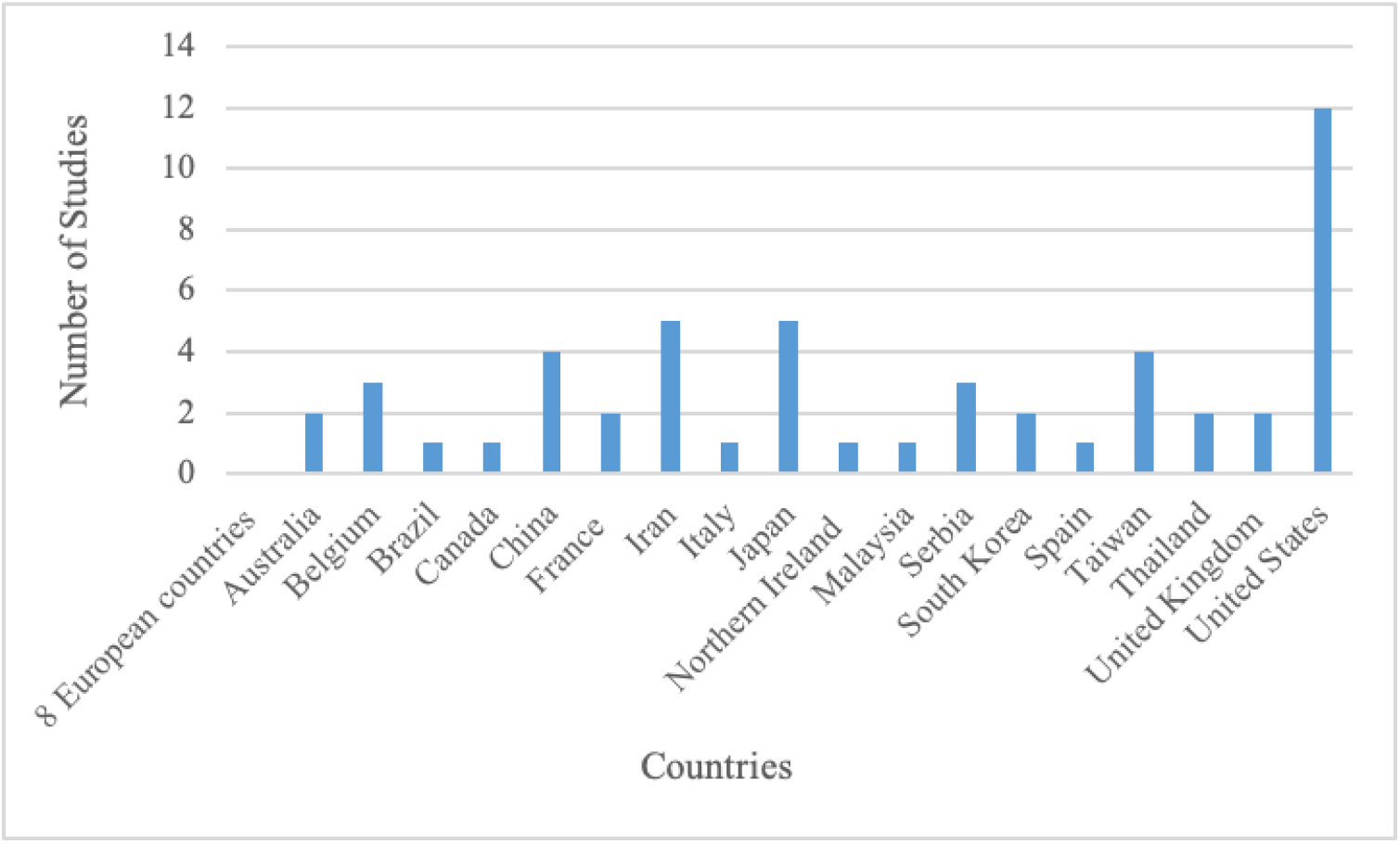
Study data countries of primary studies

#### 3.3.3 Metabolic syndrome risk factor distribution

The MetSR-F identified by the clinical definitions (See Table 1) are generally classed into invasive and non-invasive measures [65]. FBG, HDL-C, and TG are considered as invasive variables because they require the drawing of blood sample and analysis. The non-invasive risk factors are WC and BP. WC is used to identify central obesity in MetS. Other non-invasive measures used to identify central obesity in the primary studies include WtHR, BMI, and WtHtR. BP is identified by measuring systolic and diastolic blood pressure readings. In addition to the clinically identified MetSR-F, some of the studies included in this systematic review have identified and used other risk factors in the non-clinical methods such as such as age, sex, smoking habits, literacy rank, physical activity, and alcohol consumption. A single study, Lin et al. 2014 [55] collected serum samples and extracted single nucleotide polymorphisms (SNPs) associated with MetS traits for use as input parameters in their studies. The remaining 52 studies used all the five clinically defined MetSR-F in Table 1.

#### 3.3.4 Performance Metrics used in included studies

The most frequently used metrics for comparing and evaluating the predictive performance of the ML techniques in the included studies are accuracy (ACC), specificity (SPEC), sensitivity (SEN), positive predictive value (PPV), area under the curve (AUC), and negative predictive value (NPV). Table 4 below presents the definitions of each performance metric.

**Table 4.**
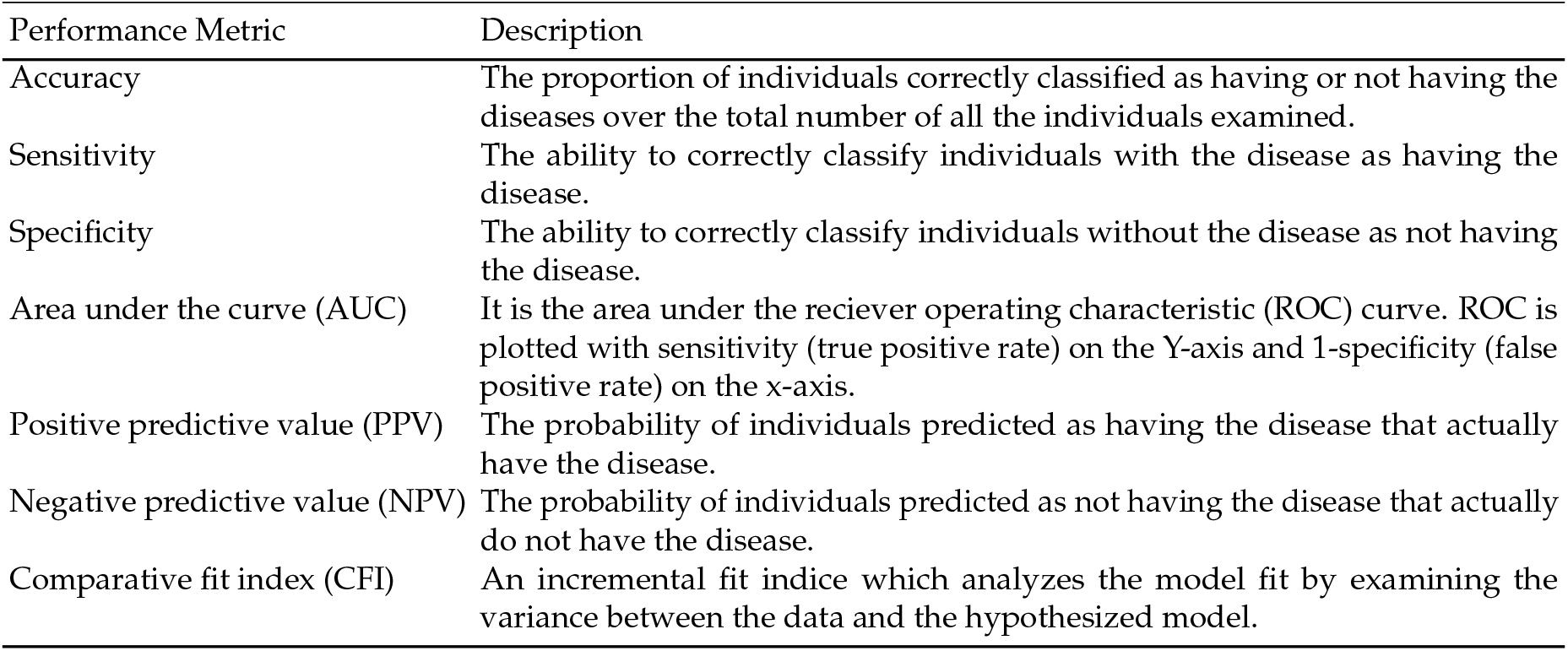
Description of performance metric measures

| Performance Metric | Description |
| --- | --- |
| Accuracy | The proportion of individuals correctly classified as having or not having the diseases over the total number of all the individuals examined. |
| Sensitivity | The ability to correctly classify individuals with the disease as having the disease. |
| Specificity | The ability to correctly classify individuals without the disease as not having the disease. |
| Area under the curve (AUC) | It is the area under the receiver operating characteristic (ROC) curve. ROC is plotted with sensitivity (true positive rate) on the Y-axis and 1-specificity (false positive rate) on the x-axis. |
| Positive predictive value (PPV) | The probability of individuals predicted as having the disease that actually have the disease. |
| Negative predictive value (NPV) | The probability of individuals predicted as not having the disease that actually do not have the disease. |
| Comparative fit index (CFI) | An incremental fit index which analyzes the model fit by examining the variance between the data and the hypothesized model. |

## 4. What is the current state of art in non-clinical methods for the diagnosis of metabolic syndrome?

The specification of a comprehensive taxonomy is a valid indication of the detailed assessment and quality of extracted data in a systematic literature review [81]. In this study, we discuss the state of art by iteratively examining and extracting relevant information from the primary studies. Although the 53 studies included in this systematic review have developed diagnosis models using various types of ML, statistical and RQ techniques for the non-clinical diagnosis of MetS, the reason why these non-clinical methods are required to support the existing clinical dichotomous diagnosis approach should be visited. The rationale for developing these alternative non-clinical diagnosis methods need to be identified in order to fully understand and analyse the state of art in the area of MetS diagnosis.

The clinical definitions of MetS shown in Table 1 are used to diagnose MetS by dichotomising the measurement values of the five clinically recognised MetSR-F (FPG, WC, HDL-C, TG, BP-SBP, DBP). MetS is considered present when the requisite number of MetSR-F that exceed certain threshold is met [4,6,82,83]. However, the MetSR-F measurement values in these definitions are more continuous than categorical and this results in loss of information on the outcome of the clinical MetS diagnosis [31,84]. Therefore, the clinical definition reduces statistical power [39,85] and patients that have MetS with only one MetSR-F present, may be excluded. Furthermore, dichotomizing the continuous MetSR scores implies that all the MetSR-F contribute equally to the diagnosis [10], however the predictive ability of some MetSR-F towards CVD is higher than others [86]. Dichotomising the continuous MetSR-F based on ad hoc thresholds could lead to mis-classification of the disorder, thereby reducing both statistical power and correlative measurement of the MetSR-F [39,85]. Also, summing up the MetSR-F into a unitary value assumes that all the risk factors contribute equally, yet some MetSR-F are known to have more importance than others [86]. Research has shown that there is a progressive relation between MetS and CVD which might be unidentified by dichotomising the MetSR-F [39,87]. Furthermore, both CVD and T2DM increase progressively as the number of MetSR-F that exceed the threshold increase, thus eliminating to apply the dichotomous definition in the diagnosis of MetS [39,87,88]. In order to solve these problems related to the dichotomous clinical diagnosis of MetS, a statistical technique called the standardised Z-score was applied to develope the continous MetS risk score(cMetSR). The cMetSR score was argued to represent and detect overall changes in the MetS dues its sensitivity to small changes in the values of the MetSR-F [32]. However the cMetSR score was limited by its over dependence on the population sample from which it was built. Subsequently, various ML techniques were then used in the primary studies to develop models for the non-clinical diagnosis of MetS as presented in Table∼∼3. Intermittently, mathematical quantification techniques were also developed and applied as quick measures of diagnosing MetS. From this systematic review, we have identified a detailed taxonomy of the ML, statistical and RQ techniques developed and applied to support the clinical diagnosis of MetS. This taxonomy and classification is presented in Fig 4.

**Figure 4.**
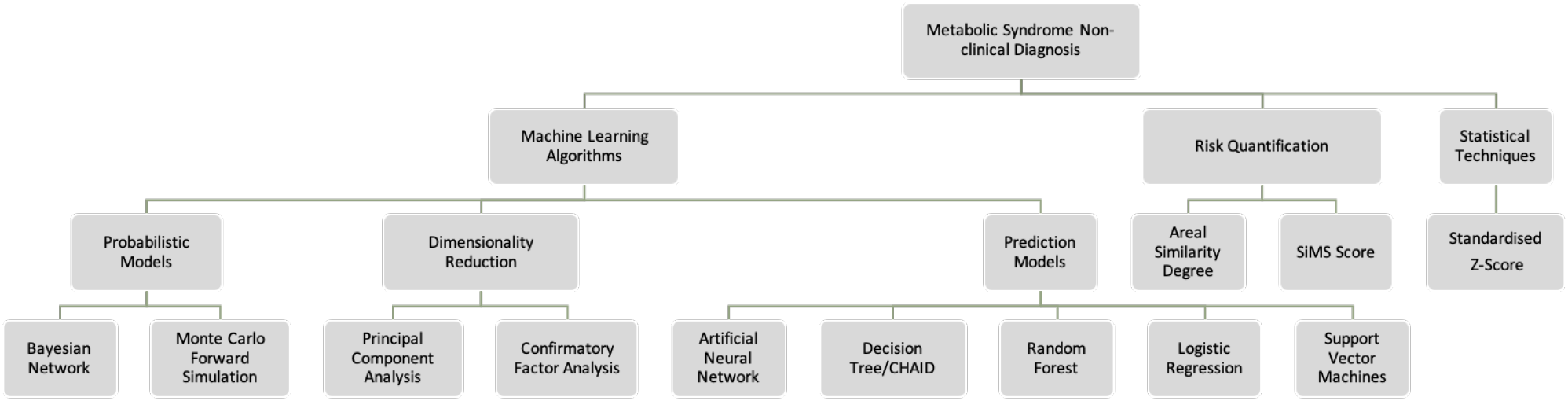
Classification and Taxonomy of non-clinical methods

### 4.1 Statistical techniques

In this section, we discuss the state of art of the 10 studies that used the Z-score.

#### 4.1.1 Z-Score

The Z-score is a statistical technique identified from the primary studies and was used to compute the cMetSR score. This score represents the presence of MetS in a population sample. The cMetSR score is computed by subtracting the sample mean from each sample value and dividing by the sample standard deviation. The higher the cMetSR score, the less favourable the MetS profile. In some studies, each of the risk factors have been regressed on age, race, and gender to account for age, race and gender-related differences in the risk factors [89]. It was widely used to calculate a cMetS in alot of studies. In Franks et al. [29], Ekelund et al. [30], and Johnson et al. [32], the cMetS was used to identify the association between physical activity energy expenditure (PAEE), aerobic fitness, physical activity and MetS. The cMetS score in [29] was highly associated with PAEE where it showed that physical inactivity may be a reason for metabolic morbidity in people with low cardiovascular fitness. Therefore, PAEE in unfit people may result in a decrease in MetSR-F such as Obesity, SBP, DBP, and increase in HDL. However, in Ekelund et al.[30] these same MetSR-F do not explain the association of MetS and physical activity. In Johnson et al.[32], there was a decrease in the risk of MetS in people who performed excersises with high PAEE. Bateman et al. [33] applied the cMetSR to conclude that increased aerobic activity resulted in a decrease in the risk of MetS and its risk factors. The cMetSR in [31]was significantly associated with increased arterial stiffness which results in CVDs. In Potteiger et al., [34], there was a significant decrease of the cMetSR score for individuals with increased PA and decreased dietary intake. Janghorbani and colleagues, [36] and Jiang et al. [37] investigated the utility of the cMetSR score computed as the standardised residuals of each MetSR-F. The cMetSR score in [36] ranged from −8.98 to 17.57 with the upper bound indicating a higher risk of MetS. Jiang et al. [37] showed that the cMetSR score was able to identify higher levels of MetS in the future based on preserved information from the history of the same participants. Thus, this suggests that cMetSR score can be applied for progressive monitoring of MetS over time. In [35], Drehmer and co-workers calculated a cMetSR score as the means of z scores of the continuous MetSR-F. The cMetSR score was used to identify if any association exists betweeen dairy consumption, and fat intake, and MetS. The CMetSR score is efficient in finding out relationships between MetSR-F and daily life style indicators. The majority of the studies reviewed in this paper showed that an active lifestyle and healthier dietary choices is inversely related with the risk of MetS.

### 4.2 Machine learning techniques

Fourty studies used different types of machine learning techniques to diagnose MetS. In this section, the methods extracted from the primary studies which include PCA, CFA, DT, DT Chaid, RF, SVM, HMM, MLR, ANN, BN, GBT and GOBAM will be described.

#### 4.2.1 Principal component analysis

Dimension reduction models are approaches for data integration that best explains the structure of datasets, and the variance both within and between variables [90]. Existing data is reduced into new variables known as components. These components best explain the difference in observations of a dataset. Two dimension reduction models were identified from the included studies: PCA and CFA. Principal component analysis (PCA) is a multivariate statistical technique that reduces the dimensions of observations in a dataset [91]. PCA is able to project the observations into a one-dimensional space that explains the variance-covariance structure of the variables in a data. The variance of the data is explained by extracting the most important information (attribute) from the observation. PCA is used to predict cumulative risk scores which enable the informative description of disease history and the development of appropriate prevention and management strategies [92].

In the studies included in this review paper, the PCA is applied to analyse the structure of the MetSR-F and the variability of their associations with MetS. Because the first principal component (PC1) of the PCA is the linear sum of measures with the maximum possible variance, researchers have used it to identify the cMetSR score [10] in MetS diagnosis. From the primary studies, 11 studies ([10],[39], [40], [41], [42], [43], [44], [45], [46], [47], [48]) used the PCA to calculate cMetSR scores.

Hillier et al. [10] created a nomogram by combining the sum of six standardised MetSR-F values weighted by the cMetSR score. The cMetSR score is the PC1 defined from the PCA which explained 50% of the variance among the MetSR-F values. They concluded that their PCA derived cMetSR score was able to predict the incidence of diabetes even in people whose FPG values were below the clinical threshold. Wijndaele et. al. in [39] and [40] also concluded that their PCA derived cMetSR score is an effective measure of MetS analysis. In [45], a continuous clinical index of cardiometabolic risk (cCICR) was constructed by first standardising each MetSR-F using the Z-score to obtain the cCICR-Z. Then, they applied PCA with orthogonal rotation on the standardised MetSR-F before finally taking the weighted sum of the two PCs (PC1 and PC2) to obtain a cCICR-PCA. Wijndaele et al. [39] found that MetS in men was higher (0*±*1.42) than women (0±1.41) and Carroll et al. [45] identified that the total variances explained by their two PCs was higher in men (61.69%) than women 60.14%. They also demonstrated that both their cCICR-Z and cCICR-PCA had higher accuracy in predicting the risks of CVD, T2DM and MetS more adequately the clinical dichotomous method.

Chang et al. [41] developed an easy-single parameter screening index called the first PC score (FPCS) by reducing only obesity (WC and BMI) and age into a single variable using PCA. This index precludes the need for using all the five MetSR-F to test for the risk of MetS in the clinical definition. Even though the FPCS was found to yield an AUC of 0.864, the computation of this parameter is inefficient for application in clinical settings. Furthermore, Agarwal and colleagues [42] investigated the use of PCA in deriving a cMetSR score as a summary of the MetSR-F and its relation to the incident of T2DM and CVD. The PC1 from their PCA model explained 33% of the total variance of the MetS risk scores. They observed that the binary cMetSR score was a better predictor of CVD than the dichotomous definition (ATP-NCEP III). The cMetSR score in [93] was calculate by summing up the scores derived by assigning points to each MetSR-F based on the size of its regression coefficient from a Cox proportional hazards model. This cMetSR was found to be useful in the prediction of CVD using MetSR-F. However,the cutoff point used is specific to only their study’s participants.

Mochizuki et al. [44] performed PCA on five MetSR-F. Their PC1 and PC2 explained 336.6 and 21.9% of the variance in the population, respectively. Even though the PCs show associations of MetS with the clinically recognised risk factors, it is still not clear if the PCA is a measure of progression of MetS and its related diseases [44]. The cMetSR score computed by Gaio and colleagues [46] using PCA was used to identify the genetic factors relevant to the risk of MetS. The first two PC scores weighted by their relative contribution in the explained variance was summed up to obtain the cMetSR score. The cMetSR score was able to explain the association of over 50% of the genetic phenotype with MetS. Wiley and colleagues, [48] developed a MetS severity score (MetSSS) also using PCA stratified by age, gender, medication and work overtime. PCA was applied to MetSR-F values standardised against clinical thresholds. The MetSSS was able to differentiate between adult with and without MetS by correctly identifying 82% of adults with MetS. However, MetSSS requires further validation in different population groups. The cMetSR was found to be useful in the prediction of CVD using MetSR-F. However,the cutoff point used is specific to only their study’s participants.

In Ayubi et. al. [47], the PCA on MetS components identified components with Eigenvalues ≥ 0.9, with 75% and 75% of the variance in males and females, respectively. Dusseault-Belanger et al.[43] examined the correlation structure of MetS using PCA and the PC1 explained 30% of the variance which was used as the cMetSR score. The cMetSR score identified MetS with a higher predictive value over the clinical definition.

#### 4.2.2 Confirmatory Factor analysis

Confirmatory factor analysis (CFA) is a statistical technique that enables the possibility of assessing existing associations between measured variables by measuring a model’s goodness of fit [94]. The CFA constructs a hypothesized model by linking various risk factors with hypothesized latent variables [94]. In the included studies, the CFA was used to develop single factor models using MetSR-F for the identification and diagnosis of MetS. CFA was used to establish the relationships between MetSR-F and MetS in order to ascertain the validity of a composite MetS risk score construct. Six studies ([49],[50], [51], [52], [53], [54]) conducted CFA on MetSR-Fs.

Gómez-Marcos and co-workers [49] developed four different cardio metabolic risk index models to diagnose MetS using CFA. All the four models consisted of TG/HDL-C ratio, HOMA-IR index, MAP and each with a different measure of central obesity - WC, WtHtR, BMI, adiposity index. The model with WC had the best MetS index with an average value of −0.022 *±*1.29 (-3.36 - 4.57) in men and the model with the BMI showed the best goodness of fit with a metabolic index of 0.0001 *±* 1.53(-3.17 -5.55) in women. The risk index was used to find associations between the MetS and physical activity (PA).

In Huo et al. [50], the CFA was used to compare two models of MetS. Both models consisted of WC, TG/HDL-C ratio, FPG, but with different measures of BP - mean arterial pressure (MAP) in Model 1 and systolic blood pressure in Model 2. WC had the highest loading both models. This reiterates the significance of central obesity in the diagnosis of MetS [6]. Model 1 showed the highest good fitness with a comparative fit index of more than 0.96 and a standardised root mean square residual of less than 0.8.

Smits and colleagues [51], CFA was applied to link new variables – adipocytokines, CT-measured intra-abdominal fat (IAF), and insulin sensitivity (SI) as underlying factors of MetS. The one factor model consisting of 6 MetSR-F - CT-measured intra-abdominal fat (IAF), insulin sensitivity (SI), SBP, DBP, TG and HDL-C had the best CFI of 0.99. However, the use of IAF and SI makes their model difficult and slow to implement in clinical settings due to the high costs and increased accessibility time of the these MetSR-F.

Gurka et al. [52], DeBoer and coworkers [53], Gurka and colleagues [38], and Musani et al. [54] all used CFA to calculate a MetS severity score (MetSSS). This MetSSS is a continuous risk score developed from a one factor CFA model consisting of the factor loadings of all the five clinically recognised MetSR-F. In Gurka et al. [52], the MetSSS revealed multiple variations in how each MetSR-F contributes to the overall MetS score based on racial/ethinic grouping. The CFA was performed on z-score standardised MetSR-F values. With an AUC ranging from 0.77, the MetSSS score was able to predict diabetes in [38]. The cMetSR score also showed a high genetic correlation with MetS in Musani et al. [54]. However, the diagnosis of MetS using the cMetSR score is heavily dependent on the score cut-off to either emphasise specificity or sensitivity. Out of the six studies that applied the CFA, three studies ([50], [51], and [52]) reported the root mean square error of approximation (RMSEA), the standardised root mean square residual (SRMR), and the comparative fit index (CFI). The CFI, SRMR, and RMSEA reported for the three studies ranged from 0.917 to 0.991, 0.134 to 0.0212, 0.125 to 0.045 respectively. CFI, RMSEA and SRMR are indices that evaluate the goodness of fit a statistical model. A model is said to have a good fit if the CFI > 0.96, the RMSEA < 0.050 and the SRMR < 0.080 [95]. These results show that all the three studies had good model fits. Only Gómez-Marcos et al. [49] reported an AUC of 89%.

#### 4.2.3 Multiple Logistic regression

Multiple Logistic regression (MLR) is a ML technique used to build prediction models for predicting values of a dependent variable from independent variables of a set of predictor values [96]. It is a linear regression variant often applied for prediction problems. The value for prediction is the probability of an event, ranging from 0 to 1. MLR also estimates risk prevalence ratios of presence of a disease and differences variable contributions to the prediction model. It works as a probabilistic view of classification.

Four studies ([71–74]) applied MLR to estimate the risk of MetS. Six other studies ([55], [56], [58], [59], [64], [77]) compared the performance of their ML models with LR. In Hosseini et al. [71], binary logistic was used to construct and validate a CMetSR score for the diagnosis of MetS in Iranian Adults. In a bottom - up, CMetSR score models were built from using two risk factors incrementally until all the five MetSR-F were included in the prediction model. The CMetSR score that includes all the 4 MetSR-F including age and gender showed the highest performance with an AUC of 95.5%. Obokata and colleagues [73], used MLR to calculate a composite risk score for MetS prediction using data from Japanese employees. The MetS risk score was used to identify the incidence of MetS in a three year followup of the population sample. A univariate LR was carried out to identify variables that would be included in the derivation of the composite risk score.

In Tan et al. 2016 [74], MLR was used to derive a cMetSR score using data collected from questionnaires. The resulting cMetSR score was able to predict MetS with an AUC of 94.2%, SEN of 90% and SPEC of 74%. Tsuo et al. [72] applied MLR to identify the relationships between the MetSR-F and the risk of having MetS in elderly people. The results show a significant positive association between all the MetSR-F and MetS.

#### 4.2.4 Artificial neural networks

Artificial Neural Network (ANN)[97] is an abstract computational structure which models nonlinear problems based on the human brain. An ANN model consists of nodes called the artificial neuron which are interconnected in a network of layers. The neuron is a simple structure capable of receiving multiple input signals via its connections but it can only output one signal. The typical ANN network constitutes of an input layer which receives data, an output layer which communicates results and sometimes with several intermediary layers (hidden layers) in-between which are used for processing [98]. The neurons between layers are linked by weighted connections which pass signals from one neuron to another. In the diagnosis of MetS where the patients are described by 6 MetSR-F (FBG, WC, HDL-C, TG, SBP, DBP), and the objective is to determine if the patient is at risk of having MetS or not, the MLP will have six inputs in the input layer and two outputs in the output layer. It is also required to define the hidden layers configuration (number of layers, number of neurons contained in each layer), their activation function and the initial synapses weights. Training entails iterative learning of synapses weights that realise the classification results with the highest accuracy. Therefore, the MLP can realize best matrix of weights that when multiplied by each of the six MetSR-F will result in the highest accurate values of risk of “MetS state”(1/0), where “MetS” = [1,0] and “No-MetS” = [0,1]. However, an ANN classifiers does not give a single class as output. For each sample, they will output a “likely answer for each class”, for example, patient x_i_’s output is probably [0.87,0.13]. This means that patient x_i_ has a 90% risk of being diagnosed MetS against a 10% risk of not having MetS.

Eight studies [55–60,62,63] applied ANN in their MetS prediction. Four studies ([58], [59], [56], [60]) developed various ANN network models trained with the back propagation (BkP) algorithm to predict MetS. BkP enables small repetitive consistent adjustments of the weights to reduce overall error in the network. The ANN models were trained and evaluated by dividing the dataset into training and testing sets respectively [58]. The ANN model developed by Chen and colleagues [58] was compared with a PC logistic regression (PCLR) model and the ANN model had a higher AUC value and SEN of 90.43% and 88.49% respectively.

Hirose et. al. compared their ANN model with a MLR model. The ANN model outperformed the MLR model with a SEN of 93% [56]. Overall, we see than the ANN model of Hirose et. al. [56] outperformed that of Chen and colleagues [58] by 4.51%. This could be attributed to the isolation of three important MetSR-F in Chen and colleagues [58] – FPG, HDL-C and TG. This information loss will result in a model with low sensitivity. In Zhao et al. [59], the BkP model was used to select single nucleotide polymorphisms (SNPs) that were associated with MetS. This model when compared with another model built with MLR model showed a higher significance in the prediction of MetS. However, obtaining SNPs may not be cost and time efficient in clinical settings.

Lin et al. [99] and Ivanovic et al. [60] explored a 3 layer network consisting of 1 to 100 neurons in the middle layer. The model with 96 hidden neurons had the highest PPV of 85.79%. However, comparison with other models was not evident in the literature and the model was built with only two MetSR-F - WtHR and BP [60]. Lin et. al. [99] compared their ANN model with a MLR model and the ANN model had a higher AUC and PPV of 93.4% and 67.5% respectively.

In Ushida et al. [57], fuzzy neural network (FNN) was applied to identify the most accurate combination of MetSR-F for the diagnosis of MetS. FNN is a hybrid combination of the fuzzy inference system and ANN [100]. Here, the fuzzy rules are extracted from the training data. A typical FNN consists of the input, membership function, rule, and output layers. Nodes at Layer 1 recieves input linguistic variables. Layer 2 computes the membership values, whose nodes represent the definition of the respective linguistic variables. Nodes at Layer 3 represent fuzzy rules. Preconditions of fuzzy rules are the links before Layer 3, and the link after Layer 3 is the consequences of fuzzy rules. Finally, Layer 4 represents the output layer. The result in [57] show that the combination of *γ*-GTP level and white blood cell (WBC) count are the most significant risk factors of MetS. However, the accuracy of the FNN model is only 68.5%. This is the least accuracy for any of the ANN models that were applied in the diagnosis of the risk of MetS.

Also, a variant of the ANN called the Auto Contractive Map (AutoCM) was used by Vigna et al.[63] to identify the complex relationships between MetS and obesity, gender, and other psycho-social risk factors. AutoCM is a technique that creates a visualization MetS and its identified risk factors in a semantic connectivity map. The map identifies complex similarities, hidden trends and nonlinear associations among attributes in a dataset. The maps are generated using the AutoCM techniques which computes the multi-dimensional association between the characteristics of each variable and all the other variables. It assigns a numerical coefficient (weight) to each link between the variables that denote the strength of their association. The links are represented as matrix connections that preserver nonlinear associations between attributes [101]. The results of the study showed that there is a strong positive relationship between the diagnosis of MetS, work- related factors and psycho-pathological variables in men and women.

Kakudi et al. [62] developed a novel ARTMAP algorithm - the Genetically-Optimized Bayesian ARTMAP (GOBAM) for the early diagnosis and risk prediction of MetS in patients who present with borderline MetSR-F measuremente. ARTMAP is an adaptive artificial system capable of solving the stability-plasticity delima which leads to catastrophic forgetting. In [62], the training sequence and parameters of the Bayesian ARTMAP (BAM) are optimised using genetic algorithm (GA). The GA is utilised to search for best combination of parameter values and sample sequence during training. Kakudi et al. [62] developed an intelligent model using GOBAM to diagnose the risk of MetS for enhancing the quality of life of patients. The predictive performance of GOBAM when compared with three other variants of ARTMAP (fuzzy ARTMAP (FAM), GA-FAM and BAM) outperformed all the ARTMAP variants with accuracy and sensitivity and ranging from 85% - 90%, and 91% - 95% respectively. However, the specificity of the GOBAM model was low with a range of 76% - 87%. There a risk quantification value generated by the GOBAM for diagnosing the risk of MetS. This value was able to identify patients with borderline measurements of MetS risk scores.

#### 4.2.5 Decision trees

Three studies ([61,64,65]) investigated the use of Decision trees (DT) to diagnose MetS and identify combinations of the MetSR-F significantly associated with its prediction. DT are trees that classify data by recursive partitioning into hierarchical or sequential structures [102]. DTs reduce the volume of data into an accurate informative summary consisting of the most important characteristics of the data. The DT consist of nodes and each node represents a decision rule that may split into two or more partition. These rules are automatically constructed and can be used for inferential decision making in clinical diagnosis. The DT model for the diagnosis of MetS for a middle-aged male will build a classification tree by identifying which of the MetSR factors should be the tree’s root node based on some selection criteria. Let’s assume the “TG” is chosen as that root node, and it has two possible values “ ≥ 1.7 mmol/L” and “< 1.7 mmol/L”. The training instances will be divided according to the level of “TG” values and this attribute will no longer be included in the training. So for each branch of “TG”, the attribute with the most relevant information must be found using the same logic. For example, for “*≥* 1.7 mmol/L”, the “HDL-C” with branches “< 1.0 mmol/L” and “> 1.0 mmol/L” could follow. This process is repeated until all the samples are classified in the same class or until all the samples have been trained. Two of the studies ([64] and [65]) proposed the use of decision trees with chi-squared automatic interaction detection (CHAID) methodology for the early detection of MetS. The chi-squared automatic interaction detection method is an algorithm used for finding patterns in datasets by merging, splitting and finally applying a user-specified stopping criteria[103].

Romero-Saldańa and colleagues used only non-invasive MetSR-F - WtHR and BP to develop a DT model. WtHR and BP were identified as the MetSR-F with the highest association to the risk of MetS in the DT model. They validated the method against the NCEP ATP III MetS definition and reported an accuracy, sensitivity and specificity of 94.2%, 91.6% and 95.7% respectively. However, Miller et al. [64] utilised all the five clinically accepted MetSR-F used to define MetS in their DT model. They reported a higher classification accuracy of 92.3% than Romero-Saldańa and colleagues. This could be attributed to their inclusion of more MetSR-F (5) which should expectedly yield a better performance decision tree model. However, noise can significantly decrease the accuracy of a DT model [104].

In [61], Worachatcheewan and colleagues proposed quantitative population-health relationship (QPHR) for the diagnosis of MetS. QPHR was developed using SVM, ANN, DT and PCA. Also [61] applied association analysis (AA) to identify the variables that repeatedly cluster together to diagnose MetS. The DT model outperformed both ANN and SVM models with an accuracy of 99% against 98% and 91% for ANN and SVM models. PCA was able classify samples as have MetS or no-MetS. The AA identifies TG+BP, BP+FPG, and TG+FG as the MetSR-F that cluster together diagnose the risk of MetS.

The gradient boosted tree (GBT) based technique was applied by Shimoda et al. [77] to develop another prediction model for the diagnosis of MetS by combining MetSR factors and psycho-social risk factors. The GBT is a variant of the gradient boosting machine developed as an ensemble of the classification and regression tree (CART) [105]. The GBT works by boosting the predictive capacity of an ensemble of weak classifiers (Trees), thereby increasing its accuracy. Compared with how RF builds its trees independently, GTB builds its trees by adding one classifier at a time. Thus, the newly added classifier is trained to improve the predictive capacity of previously trained ensemble of classifiers. For each performance score allocated to a classifier, the sum of the scores of all the classifiers is computed to obtain the final predictive score. The predictive performance of the GBT model in [77] was compared with that of MLR and RF models generated with the same dataset. The performance evaluation of the models showed that the GBT outperformed both MLR and RF models with an AUC, sensitivity and specificity of 89.3%, 87.7%, and 78% respectively. This predictive performance could be attributed to GTB’s ability to continuously correct the errors of previous trees as new trees are built up during training.

#### 4.2.6 Random forest

Three studies ([55,69,70]) applied the random forest (RF) tree algorithm to predict the presence of MetS, determine its prevalence and find significant risk factors related with the presence of MetS. Random forest is an ensemble method which combines several individual decision trees for classification and prediction [106]. RF generalises by using the bagging strategy to build each decision tree independently, thereby decreasing variance [107]. The ensemble of individual trees makes adjustment for the instability of individual trees thereby increasing the robustness of the RF method. RFs uses individual DTs as individual classifiers. However, for each tree node, a certain number of features are selected out the total number of input features. and the best split of the features divides the node. Finally, the class that is voted the most is chosen out of the trees in the forest either by regression or by classification. Nevertheless, model interpretation from the RF trees is more complex than from individual decision trees because the influence of the risk factors do not directly correspond to the risk factor’s position in the tree. Generating a multitude of DTs and then combining their predictions decreases the problem of overfitting which is usually associated with classification in DTs [108].

In the case of Lin et al. [55], they investigate the metabolic profiling changes using serum samples in MetS. First the MetS serum sample was analysed using Gas chromatography-mass spectrometry (GC-MS). Then RF models were created using metabolites from the metabolic profiling. Each MetSR-F’s level of contribution to the RF model was calculated. A proximity matrix which identifies the structure in the data was used to construct multi-dimensional scaling (MDS) plots. Similar samples have high proximity. The ACC for the RF model in [55] was 86.5% while the SEN and SPEC were 89.86% and 84.04% respectively showing significant discrimination between individuals with MetS and healthy controls.

In Worachartcheewan and colleagues, [70] data was divided into 2 subsets using PCA. The first subset is an internal dataset used for training the RF model by applying the 10-fold cross validation procedure. The second subset is an external dataset for evaluation of the RF model and FPG, WC,and BMI were the MetSR-F with the highest association to MetS according to the RF model. The importance of each MetSR-F was evaluated using the Gini index. The accuracy, sensitivity and specificity of their RF are 98.02%, 94.81% and 99.07% respectively.

Szabo et al. [69] built a RF model for predicting MetS using genetic parameters and dietary information. Their model had an ACC of 71.7%. The findings confirmed that genetic parameters are significantly associated with risk of MetS.

#### 4.2.7 Support vector machines

Two studies ([67,68]) applied Support vector machines (SVM) in the diagnosis of the risk of MetS. Karimi-Alavijeh et al. [68] explored the use of both SVM and DT to predict the risk of MetS. SVM is an algorithm that finds the optimal decision hyperplane which maximizes the separation line between data points of different classes [109]. SVM is a linear classifier that creates an optimal hyperplane which separates samples of two classes using least square regression [109,110]. It can work with mixture of both numerical and categorical data. Even though it is has accurate predictability, it is a black box technique that disallows interpretation of the classification model.

The study of Karimi and colleagues [68] explored the use of both SVM and DT to predict the risk of MetS. The SVM model proves to outperform the DT model with accuracy, sensitivity and specificity of 75.7%, 77.4% and 74.0% respectively.

Three models for predicting MetS using ANN, MLR and SVM were developed by Van Schependom et al. [67] in psychiatric patients. The three machine learning models had an accuracy, sensitivity, and specificity of between 77% - 79%, 62% - 92% and 69% - 98% respectively. They conclude that MetS can be diagnosed using less complicated ML techniques with non-invasive risk factors. However, the performance of their model is highly dependent on schizophrenic patients with decreased central obesity.

#### 4.2.8 Bayesian network

Bayesian network (BN) is a probabilistic modelling algorithm based on Bayes’ Theorem which defines the probability of an event given the occurrence of another related event. It categorizes data by monitoring the probabilities that specific features are related to specific classifications. The BN leverages on its ability to analyse results into meaningful information given an existing knowledge domain. It has the ability to handle uncertainty in complex problems. Only Miyauchi and Nishimura,[66] applied Bayesian Network (BN) modeling to connect information from specific health check-up data from Japan. Information from the BN was used to provide lifestyle advice to patients identified as being at risk of MetS. The BN model was confirmed as being a useful support tool in specific checkup and guidance system. It empowers individuals to find problems in their lifestyle and appropriate medical and health solutions easily.

#### 4.2.9 Reverse Engineering and Forward Simulation

A “Big data” analytic platform called the reverse engineering and forward simulation (REFS) was used to develop a model for the prediction of MetS using dataset generated from health insurance companies by Steinberg and colleagues [76]. REFS is big data analytic platform that is used to analyse big data sourced from multitude of sources [111]. It uses causal analytic to transparently discover the cause and effect associations in data by simulating non-parametric datasets. Learning in REFS takes place by Metropolis Monte Carlo sampling from the posterior of the model-structure distributionv[**?**]. The REFS model was able to yield interminable answers in terms of probabilities and make realistic predictions of the risk of MetS. WC was identified as the major risk factors directly associated with the risk of MetS. A decrease in WC will subsequently reduce the risk of having MetS.

#### 4.2.10 Association Rules Analysis

Huang [75] applied an association rules analysis (AA) algorithm called the data cutting and inner product (DCIP) method in order to investigate the association between MetSR-F and the risk of MetS in factory workers. DCIP partitions, sorts and carries out inner production operations on data in order to speed up the data mining process and improve computation efficiency. Fifteen association rules were generated by the AA. Medical doctors were interviewed to evaluate the efficiency of the association rules. They agreed that about 80% of the association rules derived are recognized in the medical literature to provide health guides for the diagnosis and management of MetS.

### 4.3 Risk quantification models

The areal similarity degree (ASD) was a mathematical technique proposed by Jeong and co workers, [78] for MetS risk quantification. The ASD is a similarity analysis between the MetSR-F thresholds and MetSR-F sample measurements in a weighted radar chart. The outcome of the similarity analysis is value which determines the presence or absence of MetS based on a defined cut-off value. Although the proposed model was able to diagnose MetS in individuals with borderline measurement presentations, it is sensitive to the frequency of the population sample and the positioning of the MetSR-F on the weighted radar chart.

Soldatovic and colleagues, [80] developed and evaluated the siMS score, continuous MetS score: siMSscore = 2 ∗ *Waist/Height* + *Gly*/5.6 + *Tg*/1.7 + *TAsystolic*/130 −*HDL*/1.02 or 1.28 (for male or female subjects respectively). There was a high correlation between the siMS score and the cMetSR scores derive from both Z-score and PCA. The siMS also outperformed the cMetSR scores with an AUC of 92.6%.

Yousefzadeh et al. [79] investigated the use Framingham risk score (FRS) for the prediction of MetS. The FRS is clinical tool used to asses the risk level of coronary artery disease and identifying the chance of developing any CVD in long-term [112,113]. There was a significant association between the FRS and the presence of MetS in predicting the risk of CVD in both men and women, 39.5% and 18% respectively. The odd ratio of risk of MetS was 6.7 in the high-risk FRS group (P *<*0.001).

## 5 Discussion and future guidelines

The cMetSR score, developed using Z-score, PCA and CFA, is a unitary score that has been determined to have a higher MetS risk diagnosis result than the clinical dichotomous definition [114]. A high score indicates a high MetS risk while a lower score is an indication of a less alarming risk of MetS. It is also capable of predicting the incidence of T2DM [36,45,115] and CVD [10] compared with the clinical dichotomous definition. Currently, the cMetSR score was frequently used to determine the association between MetS and other emerging risk factors. However, despite its ability to maximise statistical power [85] on the cut-off point of the MetSR-F by reducing loss of information, the cMetSR score is limited in its diagnostic capacity because all MetSR-F measurements are assumed to have an equal contribution into the diagnosis of MetS [116]. Furthermore, the cMetSR score is constrained in its application because it is a sample - specific statistical measure. This indicates that the individual score of a single patient cannot be the same in two different studies. There is also no provision to compare mean scores derived from two different studies due to differences in demographics distributions, measures of central tendency and variabilities related to the sample data. Out of the 10 studies that used statistical techniques, only five studies, ([45], [36], [38], [53], and [49]) reported AUC’s ranging from 61.3% to 89%. The six studies ([49], [50], [51], [52], [53], [54]) that used the CFA reported the goodness of fit of their models. However, there was no evidence of performance evaluation comparison in all the statistical models in the primary studies. Therefore, the predictive performance of the cMetSR score against other non-clinical methods is required in order to ascertain its efficiency. More research is therefore required to evaluate the cMetSR score against other types of MetS indexes.

The fourty studies that used machine learning techniques evaluated their methods using the performance metrics defined in Table 4. Ten studies ([61], [59], [67], [68], [74], [55], [70], [64], [65]) reported the accuracy of their models. The average accuracy performance of each technique that was used in the included studies is presented in Figure 5. ACC, SEN and SPEC values were the most frequently reported metrics by these studies. The other four studies ([58], [60], [71] and [73]) reported the AUC of their models. The graphs shows the average accuracy of 4 DT, 2 ANN, 2 MLR, 2 RF and 3 SVM techniques as reported in the 10 selected studies. The REF technique was separately used by one study each. It is clear from the graph that RF and DT were reported as the most frequently accurate techniques and the least accurate is the SVM.

**Figure 5.**
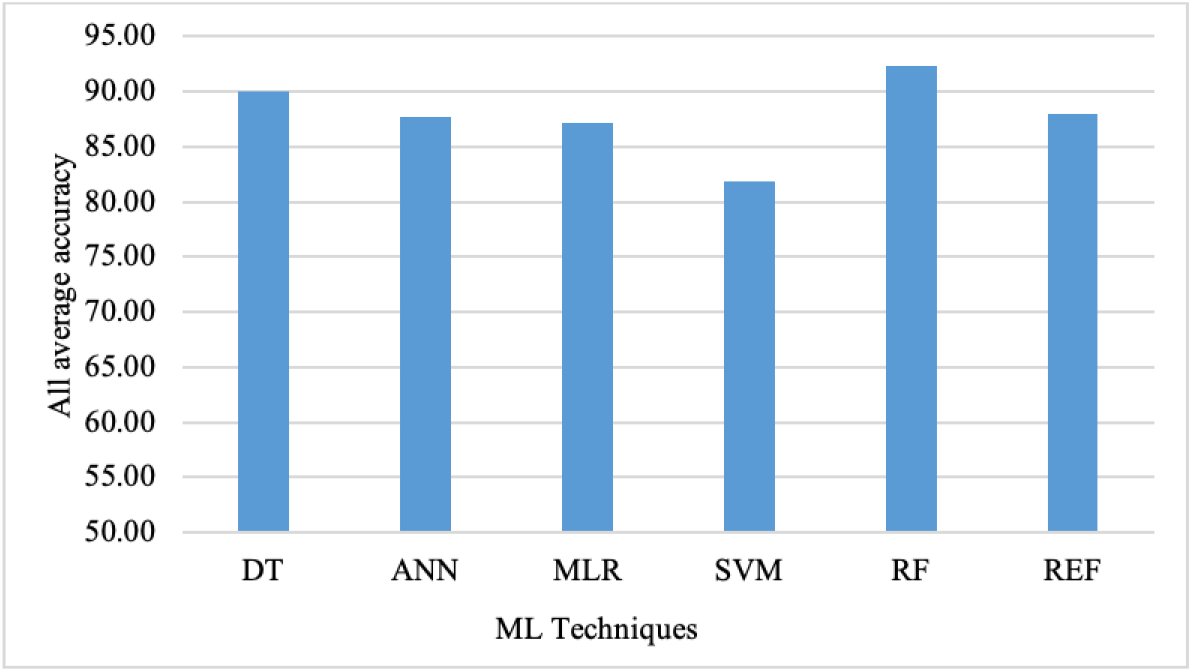
Overall average accuracy reported for each ML technique.

Most of the studies that applied statistical techniques used the PCA to develop the cMetSR score. The PCA computes cMetSR score using the principal component that explains the largest variance while maintaining the structure of the data. However, the Z-score is only a normalization technique and data preservation less evident than in the PCA. The most frequently used ML techniques are ANN, BN, and variants of DTs. The use of ANN could be ascribed to its efficiency in solving non-linear complex problems by being able to model any functional relationships and data structure. However, the BkP algorithm which is used to train the neural network, is a gradient descent technique that is characterized by getting stuck in local minima and slow convergence, limiting its application to real life domains such as the prediction of MetS [117]. Also the BPNN is depends heavily on its learning parameter settings. The use of BNs for prediction and classification problems have been successfully applied in the medical domain [118]. This could be due to its ability to handle uncertainty and integrate previous knowledge to support causal relationships. Nevertheless, performing inference creates an expensive computational burden due to the inversion of finite elements. Additionally, DTs are favored owing to their ability to generate interpretable results.

Significant conclusions can therefore be drawn from the performance measures applied to evaluate the prediction models. Consequently, the goal of ML models for predicting MetS should be to have a high predictive performance and generalization capability that enables the diagnosis of the maximum number of individuals that have or are at risk of MetS. Out of the 4 techniques mentioned, ANN, DT, SVM, and MLR, the machine models developed with ANN, DT, SVM, can be said to consistently perform better than the MLR. However, a clear winner is difficult to ascertain as the techniques show varying performances with regards to their accuracy, sensitivity and specificity in different studies. This variance could be attributed to the difference in size and dimension of the datasets. With regards to the RF, conclusions cannot be made until further studies have been conducted which compares it with other relevant ML techniques. Accordingly, more number of studies which perform comparative evaluation between various ML techniques are required in order to ascertain more generalizable models that can quickly and accurately diagnose MetS.

Oversampling methods such as SMOTE [119] to balance the training dataset and also to remove noise from the whole dataset [120] should be applied in pre-processing stages of MetS diagnosis. In addition, datasets should be cleaned using data cleaning methods such as Tomeks links [121] and Wilson’s Edited Nearest Neighbour Rule [122], to remove any overlapping that may occur with the application of oversampling methods[120]. Perveen et. al. [123] applied various sampling techniques to pre-process MetSR-F measurements in imbalance datasets before applying ML techniques to predict diabetes. Furthermore, more research using different population samples is required for better generalizability of non-clinical methods.

IThe techniques applied to develop the ML models in this systematic review assume an outcome of either being at risk or not at risk of Metabolic syndrome. This binary prediction only agrees with a correct or incorrect outcome. However, for the prediction of the risk of MetS, it is recommended that ML algorithms that also predict the probabilities of the binary outcome should be used in determining the impact of the risk of MetS in the diagnosis. This probability will aid clinicians and individuals on the best management guidelines to follow for prevention and treatment procedures. The GOBAM model has two advantages: the ability to diagnose MetS and predict the risk of MetS in patients who have MetSR -F measurements very close to the clinical threshold. It generates a probabilistic risk quantification index for assisting clinicians in the diagnosis of MetS.

Furthermore, the robustness of a ML model can also be determined based on the number of algorithms used for its comparative evaluation. For the studies in this systematic review that have conducted comparative model evaluation, it can be argued that the number of ML algorithms used for evaluation are inadequate for determining the robustness of the proposed models. From the primary studies in this category, only five studies (Worachartcheewan et al. [61], Chen et al. [58], Zhao et al.[59], Van Schependom et al. [67], Karimi-Alvijeh et al. [68], Miller et al. [64]) compared their proposed models with other existing ML techniques. Even in these studies the number of comparable models for evaluation is less than sufficient to determine the robustness of a proposed model. However in majority of the studies, appropriate procedures for evaluation, such as splitting the dataset into training, testing, and validation sets or the use of cross-validation was applied. Therefore, for proposed ML models which predict MetS, more number of existing ML models should be used for comparative evaluation for robustness and predictive generalisation.

In the case of the risk quantification techniques, no performance metrics were reported in any of the studies. Therefore, risk quantification techniques will need to be evaluated on other population samples due to their high dependency on the examination results of the population sample. Furthermore, the performance of these methods is required in order to ascertain how well they can generalised in quantifying the risk of metabolic syndrome. Majority of the primary studies are cross-sectional, making it difficult to identify the impact of the different MetSR-F on the disease outcome. More longitudinal studies could be carried out to investigate how these factors interact with each other [92]. This information may be useful in ascertaining the individual effects of each risk factor in developing future algorithms. Finally, the growth in the non-clinical approaches is encouraging with studies showing promising results. However, there is the need for further studies as follows:

1. As the cMetSR score was mostly derived for the purpose of finding associations between MetS and its related diseases (CVDs and diabetes) and other risk factors, its effectiveness for use as a tool for the diagnosis of MetS in clinical setting has not been evaluated. Therefore more studies are required to evaluate the use of cMetSR score for clinical use.
2. Research that perform comparative performance evaluation using statistical techniques with various population samples should be carried out, so as to obtain generalisable statistical models.
3. Due to imbalance in MetS datasets, preprocessing techniques such as SMOTE should be applied before deriving prediction models.
4. Studies using ML techniques should perform comparative evaluation between new models derived and other relevant ML techniques in order to ascertain the robustness of the non-clinical models.
5. Inference based ML techniques should be applied to derive non-clinical models due to their ability to present probabilistic values for MetS prediction.
6. Studies that evaluate mathematical quantification techniques using performance metrics should be conducted.

## 6 Limitation

Our analysis was limited to only studies published in the searched databases and written in English language. Secondly, due to the diverse algorithmic structures of the identified methods (Fig 4), a direct comparison between all the studies could not be carried out. For example, while ML algorithms have the ability to learn patterns from data, mathematical quantification techniques cannot learn. Thirdly, data extraction bias might have occurred because data was extracted by only one reviewer. Potentially, systematic reviews are also prone to selection bias. However, two of the reviewers independently selected the studies thereby minimizing the risk of this bias. Other limitations beyond our control such as publication bias could also be present. Often, most studies published in peer reviewed journal tend to have positive results. Nonetheless, our search scope, which spans the earliest years until January, 2020 will reduce such effect.

## 7 Conclusion

Our review shows that the three main types of non-clinical methods used for the diagnosis of MetS are ML, statistical and RQ techniques.

The ML techniques used include principal component analysis, confirmatory factor analysis, artificial neural networks, decision trees, random forests, support vector machines, multiple logistic regression, reverse engineering and forward simulation, and Bayesian networks. The artificial neural network was the most frequently used ML technique, nonetheless, highlighted proof based on performance measures shows that the random forest technique is more applicable in the development of non-clinical methods for the diagnosis of MetS. However, the random forest model tends to create large trees which makes it inefficient for quick and easy clinical application. Therefore, more alternative non-clinical methods using ML techniques should be explored to develop applications that are readily and easily available to support the clinical diagnosis of MetS in practical clinical settings.

Three mathematical quantification techniques, areal similarity degree, simScore and Framingham risk score, were also used to develop the non-clinical methods. All the models are risk quantification models which are heavily reliant on the examination results of sample participants.

This study has several implications in personalised and public health management. It provides an opportunity for researchers and health-care practitioners to gain an insight into the current trends and development of existing ML, statistical and risk quantification methods used for the diagnosis of metabolic syndrome. Indexes derived from these non-clinical methods could be used as tools which serve as quick and early preventive indicators to guide the treatment and monitoring of ongoing management of MetS and its associated diseases such as T2DM and CVD. This study sought to equip researchers and clinicians with a comprehensive analysis on the different existing classifications and application of efficient algorithms for the early diagnosis, management, and prevention of MetS and its associated diseases in health care management systems. The future guidelines of this study will guide researchers in the process of developing and advanced.

## Data Availability

Data is included in the article.

## Author Contributions

“Conceptualization, H.A.K. and C.K.L; methodology, H.A.K. and F.M.M.; validation, H.A.K., C.K.L. and F.M.M.; data curation, H.A.K.; writing-original draft preparation, H.A.K.; writing-review and editing, H.A.K. and F.M.M.; supervision, C.K.L. and F.M.M.; project administration, X.X.; funding acquisition, C.K.L. All authors have read and agreed to the published version of the manuscript.”

## Funding

“This research was supported by the Grand Challenge Grant - HTM (Wellness): GC003A-14HTM from University of Malaya and IIRG Grant (IIRG002C-19HWB) from University of Malaya.”

## Conflicts of Interest

“The authors declare no conflict of interest.” “The funders had no role in the design of the study; in the collection, analyses, or interpretation of data; in the writing of the manuscript, or in the decision to publish the results”.

## Abbreviations

**The following abbreviations are used in this manuscript**
2hPG: 2-Hour Postload Plasma Glucose
AACE: American Association of Clinical Endocrinologists
ACC: accuracy
AHA/NHLBI: American Heart Association — National Heart, Lung, and Blood Institute
AI: Artificial Intelligence
AIn: Adiposity Index
ANN: Artificial Neural Network
ART: Adaptive Resonance Theory
ARTMAP: Adaptive Resonance Theory Mapping
ASD: Areal Similarity Degree
AUC: Area Under The ROC Curve
BA: Bayesian ART
BAM: Bayesian ARTMAP
BFP: Body Fat Percentage
BMI: Body Mass Index
BN: Bayesian Network
BP: Blood Pressure
BkP: Back Propagation Algorithm
CFA: Confirmatory Factor Analysis
CFI: Comparative Fitness Index
CHAID: Chi-Squared Automatic Interaction Detection
CHOL: Total Cholesterol
CLUSTer: Cohort Study on Clustering of Lifestyle Risk Factors and Understanding its Association with Stress on Health and Wellbeing
cm: Centimeter
cMetSRS: continous MetS risk score CVD Cardiovascular Disease DALY Disability Adjusted Life Years DBP Diastolic Blood Pressure
DT: Decision Tree
EGIR: European Group for the Study of Insulin Resistance
FA: Fuzzy ART
FAM: Fuzzy ARTMAP
FN: False Negative
FP: False Positive
FPG: Fasting Plasma Glucose
FPR: False Positive Rate
FRS: Framingham risk score
FSCORE: FSCORE
GA: Genetic Algorithm
GAFAM: Genetic Algorithm Fuzzy ARTMAP
GBT: Gradient Boosted Trees
GOBAM: Genetically Optimised Bayesian ARTMAP
HC: Hip Circumference
HDL-C: High-Density Lipoprotein Cholesterol
HOMA-IR: Homeostasis Model Assessment Insulin Resistance Index
HYP: Hypertension
IAF: Intra-Abdominal Fat
IAS: International Atherosclerosis Society
IASO: International Association for the Study of Obesity
IDF: International Diabetes Federation
IGT: Impaired Glucose Tolerance
IS: Insulin Sensitivity
JDC: Japanese Diagnostic Criteria
JIS: Joint Interim Statement
kg/m^2^: Kilogram Per Square Meter
KNN: K-Nearest Neighbour
LDL-C: Low-Density Lipoprotein
MCC: Matthews Correlation Coefficient
MetS: Metabolic Syndrome
MetSSS: MetS severity score
mg/dl: Milligram Per Deciliter
ML: Machine Learning
MLR: Multiple Logistic Regression
mmHg: Millimeter Of Mercury
mmol/L: Millimoles Per Liter
MRF: Metabolic Syndrome Risk Factor
N/A: Not Available
NCD: Non-Communicable Disease
NCEP ATP III: National Cholesterol Education Program Adult Treatment Panel III
NHLBI: National Heart, Lung and Blood Institute: the American Heart Association
NPV: Negative Predictive Value
PCA: Principal Component Analysis
PCLR: Principal Component Logistic Regression
PMX: Partially Mapped Crossover
PPV: Positive Predictive Value
QMI: Quantitative Metabolic Index
QPSO: Quantum Particle Swarm Optimisation
REFS: Reverse Engineering and Forward Simulation
RF: Random Forest
RMSEA: Root Mean Square Error Of Approximation
ROC: Receiver Operating Characteristic
SBP: Systolic Blood Pressure
SEN: Sensitivity
SNPs: Single Nucleotide Polymorphisms
SPEC: Specificity
SQF: Subcutaneous Fat
SRMR: Standardised Root Mean Square Residual
STD: Standard Deviation
SVM: Support Vector Machine
T2DM: Type II Diabetes Mellitus
TG: Triglycerides True Negative
TP: True Positive
TPR: True Positive Rate
UCI: Univerisity of California Irvin
UMMC: University Malaya Medical Centre WC Waist Circumference
WHO: World Health Organisation
WHR: Waist - Hip Ratio
WHtR: Waist - Height Ratio

## Appendix A

### Appendix A.1

The appendix is an optional section that can contain details and data supplemental to the main text. For example, explanations of experimental details that would disrupt the flow of the main text, but nonetheless remain crucial to understanding and reproducing the research shown; figures of replicates for experiments of which representative data is shown in the main text can be added here if brief, or as Supplementary data. Mathematical proofs of results not central to the paper can be added as an appendix.

## Appendix B

All appendix sections must be cited in the main text. In the appendixes, Figures, Tables,etc. should be labeled starting with ‘A’, e.g., Figure A1, Figure A2, etc.

## Notes

### Competing Interest Statement

The authors have declared no competing interest.

### Author Declarations

No IRB was neccessary in this research work.

